# Development, clinical translation, and utility of a COVID-19 antibody test with qualitative and quantitative readouts

**DOI:** 10.1101/2020.09.10.20192187

**Authors:** Robert H. Bortz, Catalina Florez, Ethan Laudermilch, Ariel S. Wirchnianski, Gorka Lasso, Ryan J. Malonis, George I. Georgiev, Olivia Vergnolle, Natalia G. Herrera, Nicholas C. Morano, Sean T. Campbell, Erika P. Orner, Amanda Mengotto, M. Eugenia Dieterle, J. Maximilian Fels, Denise Haslwanter, Rohit K. Jangra, Alev Celikgil, Duncan Kimmel, James H. Lee, Margarette Mariano, Antonio Nakouzi, Jose Quiroz, Johanna Rivera, Wendy A. Szymczak, Karen Tong, Jason Barnhill, Mattias N. E. Forsell, Clas Ahlm, Daniel T. Stein, Liise-anne Pirofski, D. Yitzchak Goldstein, Scott J. Garforth, Steven C. Almo, Johanna P. Daily, Michael B. Prystowsky, James D. Faix, Amy S. Fox, Louis M. Weiss, Jonathan R. Lai, Kartik Chandran

**Author notes:** These authors made equivalent contributions (Robert H. Bortz, Catalina Florez, Ethan Laudermilch, Ariel S. Wirchnianski). Corresponding authors. (K.C.), (J.R.L.), (L.M.W.), (A.S.F.).

## Abstract

The COVID-19 global pandemic caused by severe acute respiratory syndrome coronavirus-2 (SARS-CoV-2) continues to place an immense burden on societies and healthcare systems. A key component of COVID-19 control efforts is serologic testing to determine the community prevalence of SARS-CoV-2 exposure and quantify individual immune responses to prior infection or vaccination. Here, we describe a laboratory-developed antibody test that uses readily available research-grade reagents to detect SARS-CoV-2 exposure in patient blood samples with high sensitivity and specificity. We further show that this test affords the estimation of viral spike-specific IgG titers from a single sample measurement, thereby providing a simple and scalable method to measure the strength of an individual’s immune response. The accuracy, adaptability, and cost-effectiveness of this test makes it an excellent option for clinical deployment in the ongoing COVID-19 pandemic.

## Introduction

The sudden emergence of severe acute respiratory syndrome coronavirus 2 (SARS-CoV-2), the causative agent of coronavirus disease 2019 (COVID-19), has resulted in 26.9 million cases and over 881,000 deaths worldwide to date (1–3). SARS-CoV-2 is a member of the family *Coronaviridae*, which includes six other agents known to be virulent in humans: the endemic human coronaviruses hCoV-229E, hCoV-HKU1, hCoV-NL63, and hCoV-OC43 associated with mild respiratory illness; and the zoonotic, highly virulent SARS and Middle Eastern respiratory syndrome (MERS) coronaviruses (4, 5). Infection by SARS-CoV-2 is predominantly associated with mild to moderate flu-like symptoms (6, 7). However, like the SARS and MERS coronaviruses, SARS-CoV-2 can also cause severe respiratory disease (5–7). Current COVID-19 control efforts emphasize physical distancing, molecular testing for evidence of active infection, and isolation of infected and/or symptomatic individuals and their close contacts. Antibody testing to identify individuals with prior SARS-CoV-2 infection can complement these efforts. At the community and population level, serological data can inform public health policy by uncovering spatial and temporal patterns of viral transmission. At the individual level, such testing is required to evaluate the kinetics and efficacy of the immune response to infection and vaccination. Thus, there is an urgent need for affordable and scalable antibody tests that provide both qualitative and quantitative data, ideally from single sample measurements, and that can be widely implemented.

SARS-CoV-2 entry into host cells is mediated by the viral membrane-anchored spike glycoprotein (S), which forms homotrimers decorating the viral surface (8, 9). Viral entry into human cells requires interaction of the receptor-binding domain (RBD) of the spike protein with its host-cell receptor, angiotensin-converting enzyme 2 (ACE2), which is expressed in a variety of tissues, including the lung, small intestine, kidney, heart, and testes (10–12). The spike protein is also a major target of the humoral immune response, and spike-specific antibodies that block viral entry into cells (neutralization) can afford protection against severe disease (13, 14). A number of studies have shown that convalescent patient sera contain high levels of SARS-CoV-2 spike-specific IgA, IgM and IgG antibodies with significant virus-neutralizing activity (15–18). In addition the spike protein’s sequence divergence from those of the widely circulating endemic hCoVs (<30% sequence similarity of the *S* gene at the amino acid level (19)) make it an ideal antigen to detect and measure SARS-CoV-2 seroconversion.

Here we describe a highly sensitive and specific ELISA-based test for SARS-CoV-2 exposure that was developed and clinically translated at the height of the COVID-19 pandemic in New York City in March-April 2020. The test employs a purified, recombinant SARS-CoV-2 spike protein ectodomain and readily available, research-grade laboratory reagents to detect spike-specific IgG and IgA antibodies in human sera or plasma. We show that the IgG test affords not only for the qualitative assessment of SARS-CoV-2 exposure with high sensitivity and specificity, but also the accurate determination of spike-specific IgG titers from a single sample measurement.

## Results

### Development of an ELISA to detect SARS-CoV-2 spike-specific IgG and IgA in COVID-19 convalescent sera

Available serological assays for SARS-CoV-2 have used antigens derived from the spike and/or nucleocapsid proteins, the predominant targets of the humoral response to natural infection (8, 17, 18, 20, 21). Further, many spike-specific assays have employed truncated forms of the spike (e.g., the S1 subunit or the RBD) as the target antigen (17, 22, 23), in part because the full-length spike can be challenging to produce at scale. Here, we focused on the full-length spike ectodomain as our assay antigen so as to sample antibodies that recognize all parts of the spike protein (20). Accordingly, we produced a recently described recombinant spike ectodomain protein bearing stabilizing mutations (9). Optimized expression and purification protocols resulted in yields of 20-35 mg/L of homogenous, structurally well-defined spike trimers from transiently transfected ExpiCHO-S^TM^ cell cultures (24).

We examined the capacity of this trimeric spike protein to specifically capture antibodies in convalescent sera from healthy individuals with prior SARS-CoV-2 infection. Spike protein-coated ELISA plates were incubated with serial dilutions of serum, and bound antibodies were detected and measured with a human IgG-specific secondary antibody conjugated to horseradish peroxidase (HRP). Varying levels of spike-specific IgG were detected in convalescent sera but not in a pre-COVID control serum (Fig 1a).

Although most efforts to characterize the SARS-CoV-2 humoral immune response have focused on IgG, multiple reports suggest that IgA may also provide a sensitive marker for SARS-CoV-2 exposure (15, 19, 25). Accordingly, we used the assay format described above, but with a human IgA-specific secondary conjugate, to detect and quantify spike-specific IgA in the same serum samples. IgA was consistently detected in these samples and was present at levels concordant with those of IgG (Fig 1b).

**Fig 1:**
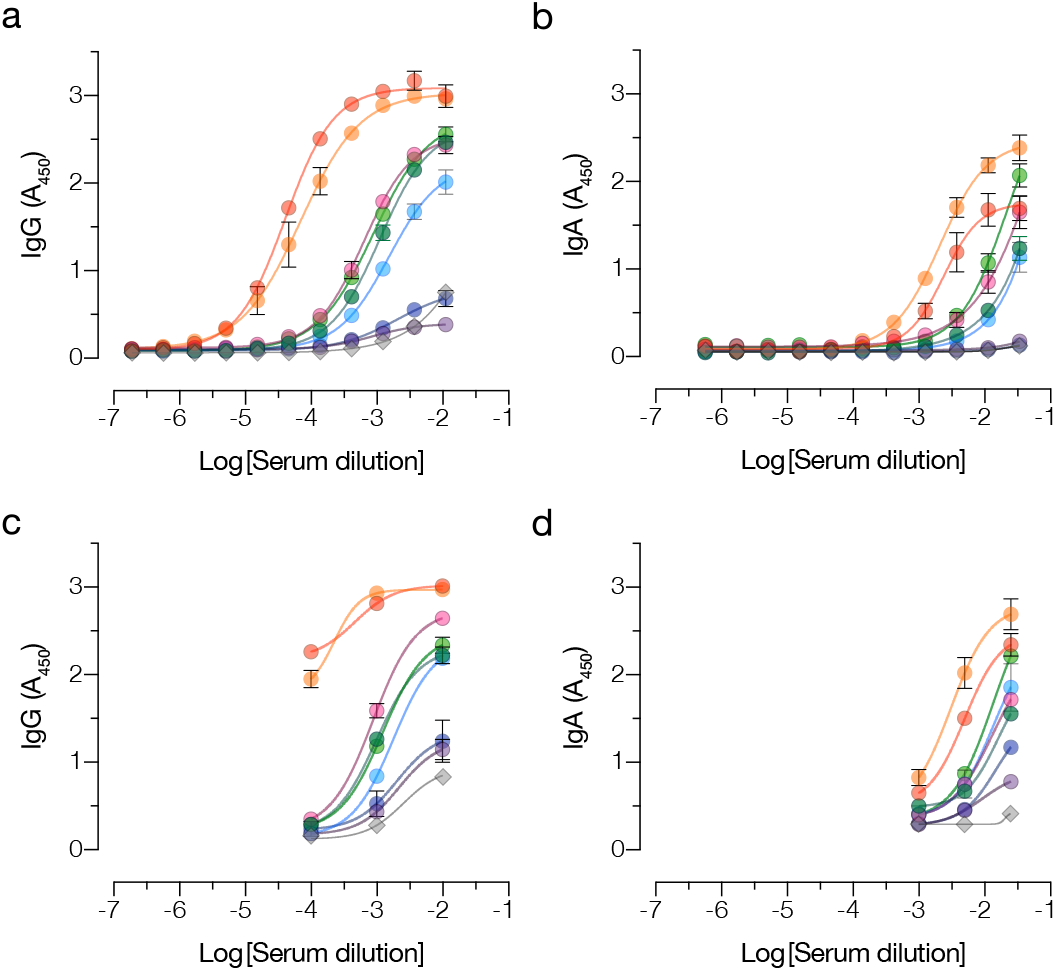
ELISA to detect and measure SARS-CoV-2 spike-specific IgG and IgA in COVID-19 convalescent sera. Serially diluted convalescent patient sera (colored circles) and a pre-2020 negative control (gray diamonds) were added to recombinant SARS-CoV-2 spike protein-coated ELISA plates. Captured IgG (a) and IgA (b) were detected using Ig class-specific secondary antibody-HRP conjugates. Absorbance (A_450_) values were fitted to a sigmoidal curve. Samples were re-analyzed at three dilutions that best characterized the extent of the antibody reactivity for IgG (c) and IgA (d). Averages +/− SD are shown, n=4 from two independent experiments. SD values smaller than the height of the symbols are not shown.

### Definition of optimal single dilutions and corresponding diagnostic thresholds for the S-specific IgG and IgA ELISAs

To develop the assay into a clinical laboratory test, we sought to identify a serum dilution which could provide a single threshold for reliably detecting spike-specific antibodies. Accordingly, we examined three sample dilutions each for IgG and IgA in an ELISA (Fig 1c, d), which were selected from full response curves (Fig 1a, b). Using this simplified three-dilution ELISA, we analyzed a large panel of sera from COVID-19 convalescent donors (Conv; presumptively seropositive) and archival pre-COVID sera (Ctrl; presumptively seronegative) (Table 1) for both IgG (Fig 2a, b) and IgA (Fig 2d, e).

Donors in the Conv cohort (n=197) were initially selected to identify potential COVID-19 convalescent plasma donors according to New York Blood Center donor guidelines at the time of screening. Specifically, they had confirmed prior infection (positive RT-qPCR for SARS-CoV-2 RNA) and had been asymptomatic for at least 14 days at the time of sampling (median 28 days post symptoms and 24 days post diagnosis). The Ctrl cohort was a set of patient serum samples collected at Montefiore Medical Center between 2008-2019 (Ctrl-Pre-2020; n=171) and in Jan 2020 (Ctrl-Jan 2020; n=45), prior to identification of the first COVID-19 cases in the greater New York City area in late February 2020 (26) (Table 1).

To assess assay reproducibility, the Ctrl and Conv samples were analyzed in two independent experiments conducted by different researchers. The average absorbances (A_450_) from the independent experiments were found to be highly correlated for both IgG (Fig 2c) and IgA (Fig 2f).

The results from the presumptive seropositive and seronegative cohorts were analyzed using receiver-operator characteristic (ROC) curves to determine assay sensitivity and specificity at each candidate threshold value (Fig 3a and Table 2). For both IgG and IgA, the area under the curve (AUC) was found to be essentially equivalent at the lower two serum dilutions but greater than that at the highest serum dilution. To conserve clinical samples for additional laboratory tests, we selected the intermediate dilution (1/1,000 for IgG and 1/200 for IgA) for further analysis.

**Table 1:** Cohorts employed in this study.

| Sample cohort | N | Gender (M/F/Unk<sup>a</sup>) | Median age [IQR]<sup>b</sup> | Median days post-Sx [IQR] | Median days post-Dx [IQR] | Notes |
| --- | --- | --- | --- | --- | --- | --- |
| Convalescent | 197 | 126/64/7 | 42 [32–54] | 28 [24–31] | 24 [20–27] | Mild disease, no O<sub>2</sub> support |
| Hospitalized | 27 | 19/8 | 65 [55–73] | 6 [1.5–7]<sup>c</sup><br>13 [10–16]<sup>d</sup> | 0<sup>c</sup><br>8 [7–9]<sup>d</sup> | Moderate to severe disease |
| Control Jan 2020 | 45 | 13/32 | 54 [39–61] | N/A | N/A | Samples collected 1/28-1/30/2020 |
| Ctrl Pre-COVID | 171 | 53/103/15 | 56 [49–62] | N/A | N/A | Samples collected 2007–2017 |
| Human CoV (Sweden) | 17 | 9/8 | 66 [46–72] | Unk | 130 [31–221] | Pre-COVID \| Swab-positive for OC43/ HKU1, 229E, or NL63 |
| Human CoV (MMC) | 5 | 3/2 | 32 [24–37] | Unk | Unk | Samples collected 1/1-4/27/20 \| Swab-positive for OC43/ HKU1, 229E, or NL63 |

<sup>a</sup> Male/Female/Unknown
<sup>b</sup> Interquartile range
<sup>c</sup> Samples collected 0–1 days post-hospitalization
<sup>d</sup> Samples collected 6–10 days post-hospitalization

**Fig 2:**
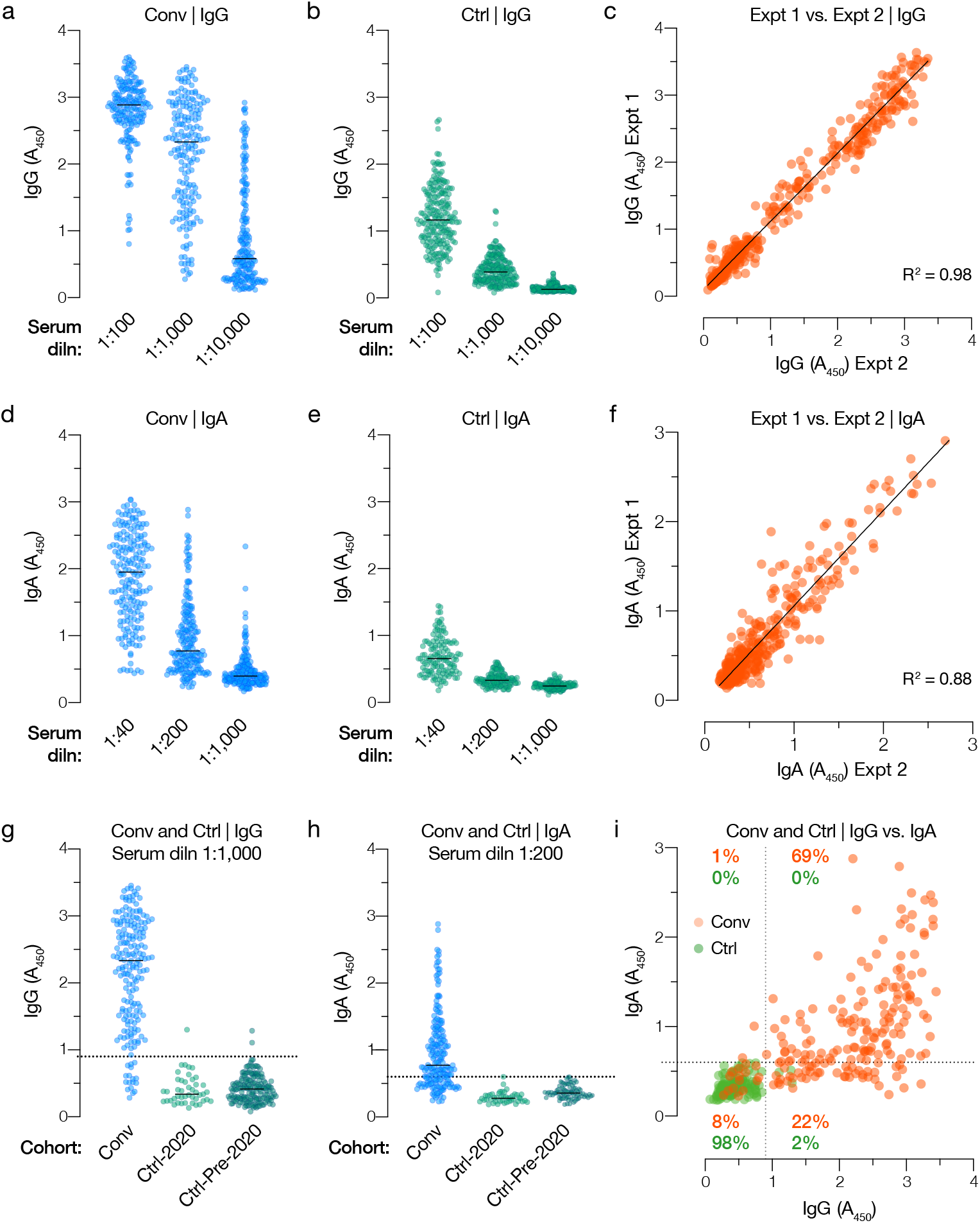
Analysis of spike-specific IgG and IgA reactivity in convalescent and control cohorts. Spike-specific IgG (a-b) and IgA (d-e) responses at the indicated serum dilutions were determined for convalescent (Conv, n=197) and control (Ctrl, n=216) cohorts. (c, f) Inter-assay reproducibility of independent IgG and IgA assays at serum dilutions of 1:1,000 and 1:200, respectively, was assessed by a linear regression analysis. (g-h) Spike-specific IgG and IgA reactivity for Conv and Ctrl cohorts at the selected test dilution (1:1,000 and 1:200 serum dilutions, respectively). Diagnostic thresholds for IgG and IgA tests are shown as dotted lines (A_450_ values of 0.90 and 0.60, respectively). Ctrl cohort is separated into Pre-2020 (n=171) and Jan 2020 groups (n=45). (i) IgG and IgA reactivities of each sample in the Conv (orange circles) and Ctrl (green circles). Respective diagnostic thresholds are indicated as dotted lines. Percentages reflect the proportion of Ctrl and Conv sera in each quadrant.

To identify the diagnostic threshold that maximized assay sensitivity and specificity at this dilution while minimizing the number of clinically harmful false positives, we selected a point on each ROC curve corresponding to a specificity of ~99%, thereby obtaining threshold A_450_ values of 0.90 and 0.60, respectively, for IgG and IgA. These threshold values were at or near the maximal point of the curve comparing the sum of sensitivity and specificity against each candidate threshold, indicating near-optimal assay performance (Fig 3b, c). Reanalysis of the datasets at these thresholds yielded sensitivities of 91% and 70% (Fig 3b, c), respectively, and a specificity of ~99% for the IgG and IgA tests (Fig 2g, h, Tables 3–4).

Given the lower sensitivity of the IgA test relative to the IgG, we determined the relationship between the test results for each patient in the Conv and Ctrl cohorts (Fig 2i). Although IgG positivity correlated with that of IgA, especially for the strongly positive sera, a considerable proportion (22%) of the IgG-positive Conv sera were negative for IgA. The converse was not true—only 1% of Conv samples were positive for IgA but negative for IgG. These findings are consistent with emerging evidence that serum IgA wanes more rapidly than IgG in COVID-19 convalescent patients (27). We conclude that IgG provides a more sensitive probe of SARS-CoV-2 exposure in convalescent patient sera than does IgA, at least when the spike protein is used as the capture antigen.

### Prior exposure to endemic human coronaviruses is not associated with false positives

The high seroprevalence of endemic human coronaviruses (hCoVs; >90% of adults over 50 years old) (28, 29) and the low positivity rates of the archival Ctrl specimens in the spike IgG and IgA tests strongly suggested that both tests specifically detect the divergent SARS-CoV-2 spike protein. To further address SARS-CoV-2 assay specificity, we tested 22 pre-COVID-19-pandemic serum samples from individuals who had RT-qPCR-confirmed infection with human alphacoronaviruses (229E, NL63) or betacoronaviruses (OC43, HKU1) for anti-S IgG and IgA by ELISA (Fig 4). In agreement with the molecular test results, we observed that most of these samples contained reactive IgG and/or IgA against the recombinant spike proteins of the hCoVs 229E and HKU1 but not the emerging betacoronavirus, MERS. However, none were positive in our IgG and IgA tests. Thus, both tests are highly specific for SARS-CoV-2 and unlikely to engender false positives due to prior patient exposure to circulating hCoVs.

**Fig 3:**
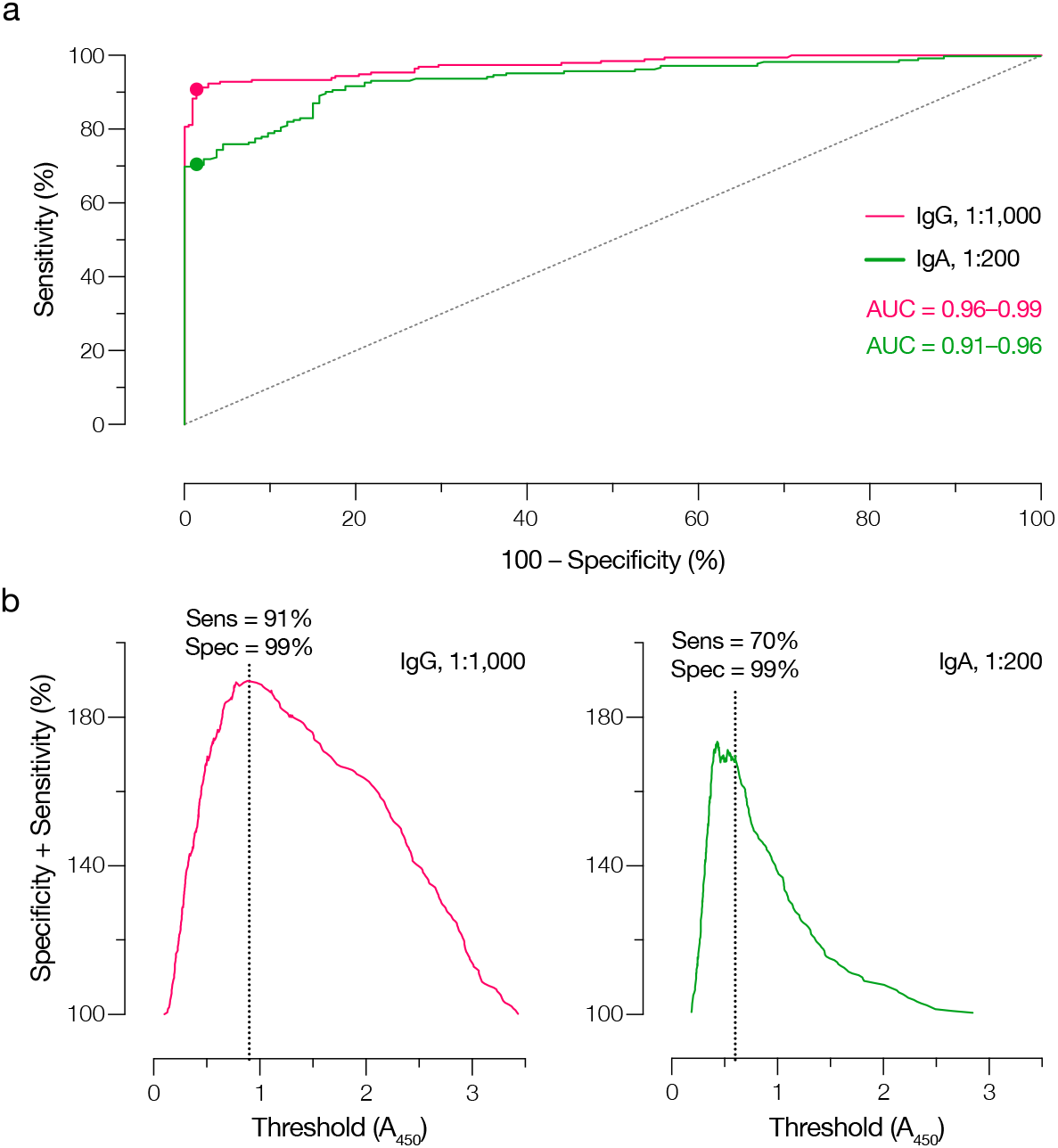
Selection of diagnostic thresholds for IgG and IgA tests using receiver-operator curve (ROC) analysis. **(a)** ROC analyses for the IgG and IgA tests. AUC, area under the curve. Filled circles indicate the point on each ROC that corresponds to the selected diagnostic threshold. **(b)** The sum of assay sensitivity and specificity for each candidate diagnostic threshold was extracted from the ROCs for the IgG and IgA tests. Dotted lines indicate the selected thresholds (A_450_ of 0.90 and 0.60 for IgG and IgA, respectively).

**Table 2:** Receiver-operator (ROC) curve analysis of SARS-CoV-2 spike-specific IgG and IgA antibody tests at three different dilutions of control and convalescent antisera.

| Antibody class | Serum dilution | Positives<sup>a</sup> (N) | Negatives<sup>b</sup> (N) | AUC<sup>c</sup> [-/+ 95% CI]<sup>d</sup> | Sens. at 99% spec.<sup>e</sup> [-/+ 95% CI] | P value |
| --- | --- | --- | --- | --- | --- | --- |
| IgG | 1:100 | 197 | 216 | 0.96–0.99 | 75–86 | <0.0001 |
| IgG | 1:1,000 | 197 | 216 | 0.96–0.99 | 83–92 | <0.0001 |
| IgG | 1:10,000 | 197 | 216 | 0.95–0.98 | 65–78 | <0.0001 |
| IgA | 1:40 | 197 | 133 | 0.89–0.95 | 64–76 | <0.0001 |
| IgA | 1:200 | 197 | 133 | 0.91–0.96 | 63–76 | <0.0001 |
| IgA | 1:1,000 | 197 | 133 | 0.88–0.94 | 45–59 | <0.0001 |

<sup>a</sup> Convalescent donor cohort (Conv)
<sup>b</sup> Pre-COVID cohort (Ctrl)
<sup>c</sup> Area under the curve
<sup>d</sup> 95% confidence intervals
<sup>e</sup> Test sensitivity at 99% specificity

**Table 3:** Results of SARS-CoV-2 spike-specific IgG test

| Sample cohort<sup>a</sup> | Positive (N) | Negative (N) | Total (N) | Positive (%) | Negative (%) |
| --- | --- | --- | --- | --- | --- |
| Conv (>d15 Sx) | 180 | 17 | 197 | 91% | 9% |
| Ctrl-2020 (Jan 2020) | 1 | 44 | 45 | 2% | 98% |
| Ctrl-Pre (Pre-COVID) | 2 | 169 | 171 | 1% | 99% |
| hCoV | 0 | 22 | 22 | 0% | 100% |
| All controls | 3 | 235 | 238 | 1% | 99% |

<sup>a</sup> Samples analyzed at 1/1,000 serum dilution

**Table 4:** Results of SARS-CoV-2 spike-specific IgA test

| Sample cohort<sup>a</sup> | Positive (N) | Negative (N) | Total (N) | Positive (%) | Negative (%) |
| --- | --- | --- | --- | --- | --- |
| Conv (>d15 Sx) | 138 | 59 | 197 | 70% | 30% |
| Ctrl-2020 (Jan 2020) | 0 | 45 | 45 | 0% | 100% |
| Ctrl-Pre (Pre-COVID) | 1 | 87 | 88 | 1% | 99% |
| hCoV | 0 | 22 | 22 | 0% | 100% |
| All controls | 1 | 154 | 155 | 1% | 99% |

<sup>a</sup>Samples analyzed at 1/200 serum dilution

### Test performance in a cohort of hospitalized COVID-19 patients

We next assessed the capacity of the IgG and IgA tests to detect SARS-CoV-2 exposure in a hospitalized COVID-19 cohort. The spike-specific IgG and IgA reactivities of blood drawn from each patient immediately (days 0-1; early samples) after hospital admission or after 6-10 days (late samples) were determined. Most of the early samples were negative for both IgG and IgA; in contrast, most of the late samples were positive for both (Fig 5a, c), indicating that most of the patients (but not all) developed a detectable antibody response to SARS-CoV-2 over the first 6-10 days of their hospitalization. Because the interval between symptom onset and date of hospitalization varied, we replotted the above data to observe the development of the spike-specific antibody response post-symptom onset. Both IgG and IgA were readily detectable by day eight and a majority of the patients were IgG and/or IgA-positive by day 14 post-symptom onset (Fig 5b, d).

### Cross-validation with New York State Department of Health SARS-CoV-2 antibody test

The Wadsworth Center (WC), the public health laboratory of the New York State Department of Health (NYSDOH) developed a microsphere-based test for SARS-CoV-2 spike-specific antibodies that has been extensively employed to date (30, 31). To cross-validate our IgG antibody test, we provided WC with ten Conv samples and two Ctrl samples for analysis in their assay. WC was blinded to all samples. Despite the considerable differences in the platforms and diagnostic thresholds employed by the WC and Einstein spike-based IgG tests, there was complete agreement between them (Fig 6a).

**Fig 4:**
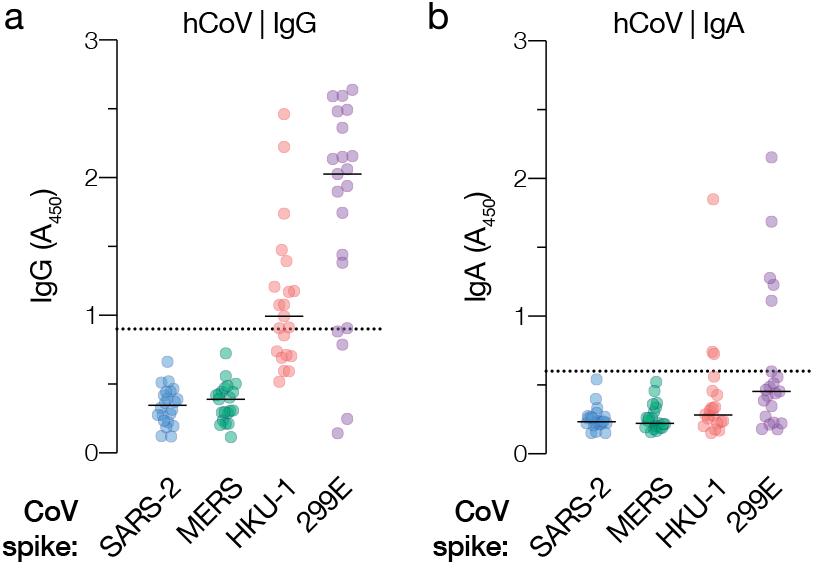
Specificity of IgG and IgA tests for SARS-CoV-2 vs. commonly circulating human coronaviruses. Serum samples from two cohorts of COVID-19-negative patients with RT-qPCR-confirmed exposure to one or more commonly circulating human coronavirus (hCoV) were analyzed in the SARS-CoV-2 IgG **(a)** and IgA **(b)** tests (SARS-2) and for reactivity against recombinant spike proteins from the indicated alpha- and betacoronaviruses. Dotted lines indicate diagnostic thresholds for the SARS-CoV-2 antibody tests only. Data from at least 2 independent experiments (n=4-8) are shown.

**Fig 5:**
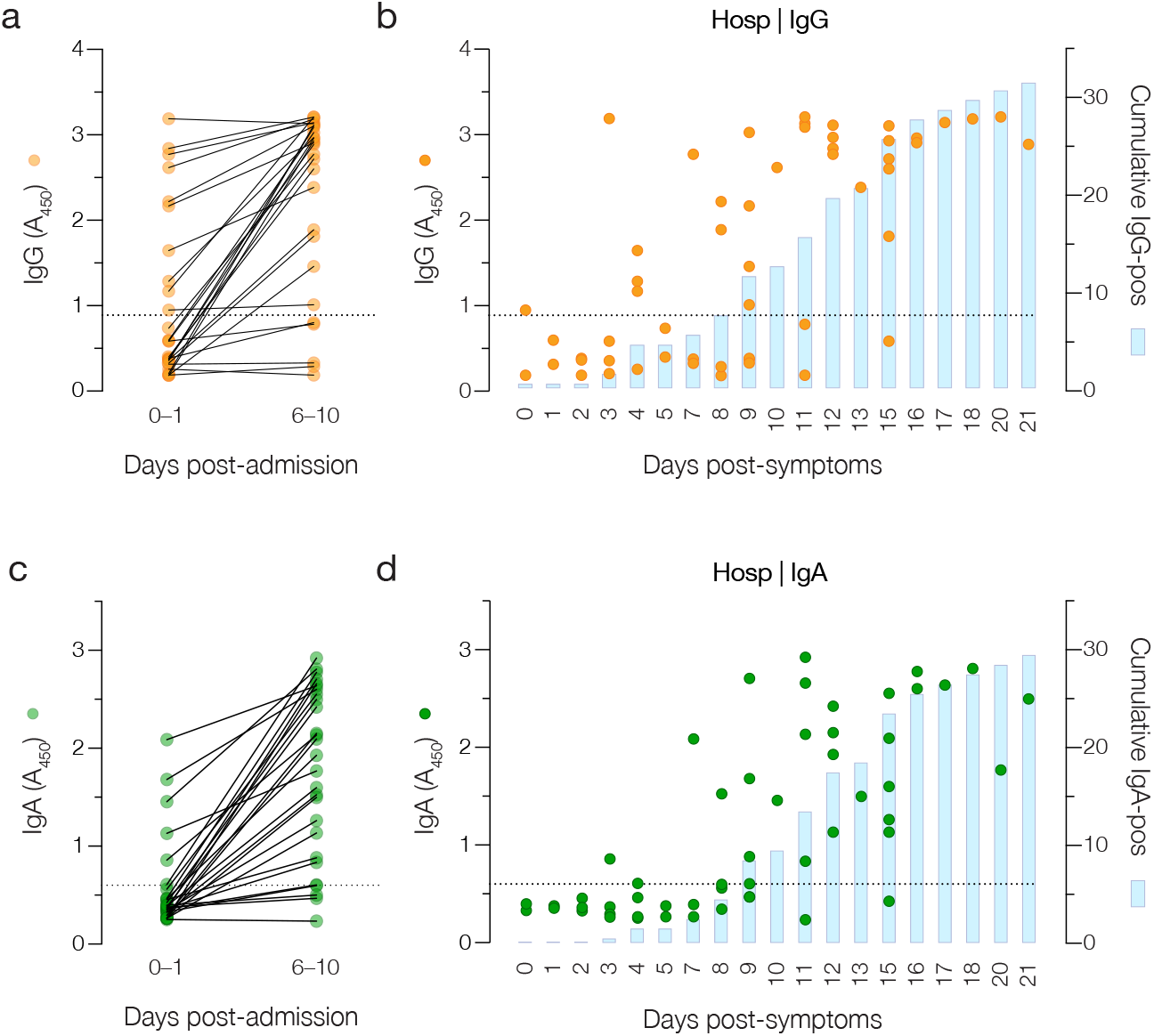
Longitudinal analysis of hospitalized patients at two early time points with the IgG and IgA tests. Serum samples from patients at two time points following hospitalization were analyzed for spike-specific IgG **(a)** and IgA **(c)**. Individual patient samples (circles) and cumulative positive results (blue bars) are graphed as a function of days post-symptom onset (self-reported) for IgG **(b)** and IgA **(d)**. Data from two independent experiments (n=4) are shown.

### Clinical translation

All of the above results presented were generated by manual experimentation in research laboratories at Albert Einstein College of Medicine. To leverage our IgG assay for high-throughput clinical testing, we translated it to a robotic platform in a CLIA-certified laboratory at Montefiore Medical Center (MMC). A larger subset of Ctrl and Conv samples (102 in total) were tested on the MMC platform. The MMC laboratory was blinded to the identities of all samples. A high level of agreement (96%) between the platforms was observed, with three of four discordant results occurring in samples with values at or near the diagnostic threshold (i.e., borderline samples) (Fig 6b). This clinically translated test formed the basis of an NYSDOH application for a laboratory developed test (LDT) by MMC.

### IgG test affords quantitation of anti-spike antibodies from a single measurement

Having established and clinically translated the IgG ELISA for use as a diagnostic test, we investigated if it could also be used to determine the magnitude of the spike-specific antibody response in patient samples. We first generated serum titration curves (e.g., Fig 1) to determine spike IgG endpoint titers for the entire Conv cohort (n= 197). Next, we compared these titers to the independently measured A_450_ values for the same samples at 1/1,000 dilution (Fig 2a) and observed a nonlinear relationship (Fig 7a). Accordingly, we modeled this relationship through a nonlinear regression analysis by fitting a sigmoidal function using the least-squares method.

**Fig 6:**
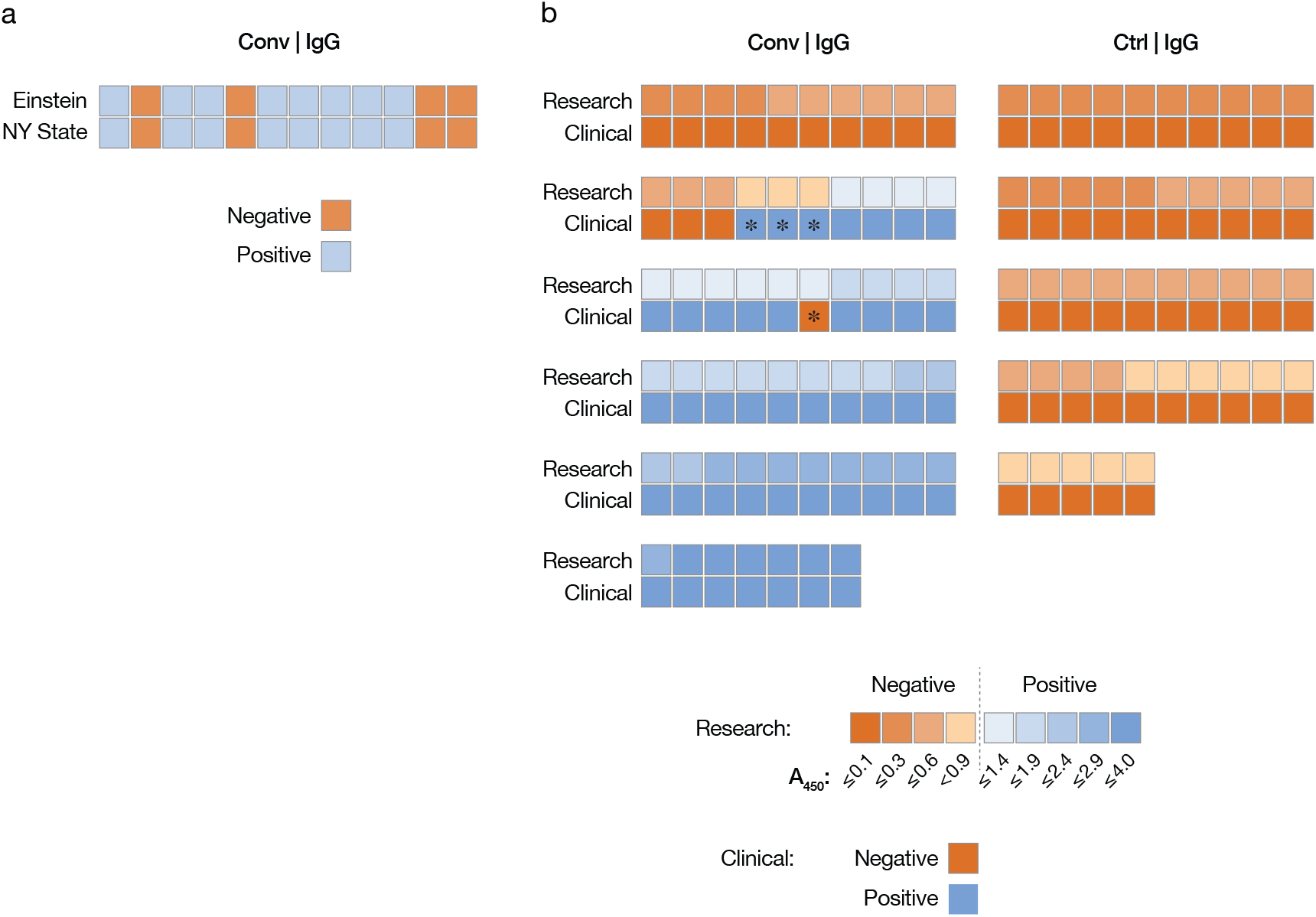
Clinical translation of IgG test and cross-validation with New York State Department of Health test. Heat maps comparing results from a subset of Conv and Ctrl samples obtained with the manual IgG test performed in the laboratory at Einstein (Research) (a-b) to those obtained with the Wadsworth Center microsphere immunoassay (NY State) **(a)** and the clinically translated test performed in the CLIA-certified laboratory at MMC (Clinical) **(b)**. Asterisks denote discrepant results.

Our model fit the experimental data well (Fig 7a; R^2^ = 0.88), suggesting that it could be used to infer spike-specific IgG titers from the single measurement performed for the diagnostic test. We further employed a ten-fold cross-validation method to evaluate the predictive utility of the model (see Materials and Methods for details). Our model could accurately predict the experimental IgG titer of a convalescent serum sample based solely on a single A_450_ measurement (R^2^ = 0.81 ± 0.02) (Fig 7b).

**Fig 7:**
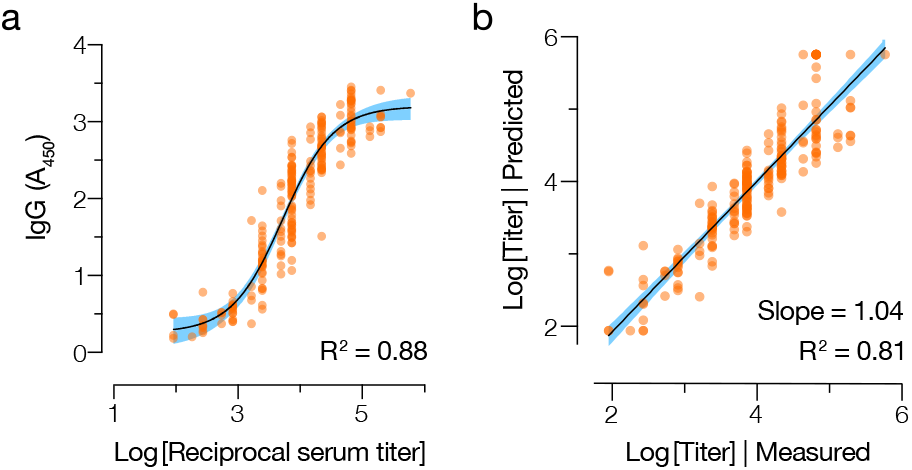
IgG test affords quantitative assessment of serum IgG from a single measurement. **(a)** The relationship between the log-transformed readout value (A_450_ at 1/1,000 serum dilution) in the IgG antibody test and the endpoint IgG titer (determined from full ELISA curves), for each serum sample in the Conv cohort. Data were fit to a sigmoidal function through a nonlinear regression analysis. **(b)** A ten-fold cross-validation method was used to evaluate the predictive utility of this model. For each serum sample, the experimentally determined endpoint IgG titer was compared to that predicted from a single measurement with the antibody test using linear regression analysis. Shaded blue areas represent the 95% confidence intervals for the curve fits.

## Discussion

As the COVID-19 pandemic continues, there remains a need for non-proprietary and scalable diagnostic antibody tests for monitoring populations that are vulnerable to SARS-CoV-2 and to gauge exposure at a population-wide level. High-throughput assays for quantitative serology are also urgently needed to support the development and global deployment of COVID-19 vaccines and convalescent plasma-based therapeutics. Here, we describe, validate, and clinically translate a simple, high-performance ELISA-based test for SARS-CoV-2 spike-specific IgG, developed at the height of the COVID-19 pandemic in New York City in March – April 2020. We also explore the utility of a highly specific IgA-based test for SARS-CoV-2 exposure. Finally, we demonstrate that our test can accurately quantitate SARS-CoV-2 spike-specific IgG in clinical samples from a single measurement.

The spike protein is a major target of the human antibody response to natural coronavirus infection and has key advantages as a capture antigen in serologic assays. First, anti-spike, but not anti-nucleocapsid antibody titers in convalescent sera correlate with neutralizing activity (17, 18, 32),decreased disease and viral load in animal models (13, 14, 33–37) and survival following SARS-CoV-2 infection (13, 14). Second, the spike gene has the most divergent protein-coding sequence among the coronaviruses. The spike protein is thus the least likely to engender false positives due to antibodies arising from endemic hCoV exposure (Grifoni et al. 2020; Okba et al. 2020). Despite these potential advantages, the nucleocapsid protein has long been favored over spike in coronavirus serologic assays, in part because it can be readily expressed at high levels without compromising conformation or immunogenicity (38). By contrast, pre-fusion trimers of the larger, more complex, and heavily glycosylated spike protein are more challenging to produce at scale (39). A number of spike-specific antibody tests have relied on individual spike subunits (typically, the highly immunogenic *N*–terminal subunit S1) (22, 40) or on truncated protein fragments (typically, the receptor-binding domain [RBD]) (17). We optimized the production and purification of a stabilized, full-length spike ectodomain described by Wrapp and colleagues (9) and showed that these scaled-up preparations largely consisted of homogenous pre-fusion trimers (24). We leveraged these large, biochemically well-defined preparations to develop a scalable serologic assay for SARSCoV-2 that could comprehensively sample the antibody response to its spike protein.

We initially sought to establish a qualitative antibody test based on a standard ELISA format. We showed that analysis at three serum dilutions could corroborate the results of full antibody titration curves for IgG and IgA (Fig 1). Further analysis of large convalescent, pre-COVID control, and hCoV-exposed control cohorts at these three serum dilutions allowed us to identify optimal single dilutions and diagnostic thresholds for both tests. At the selected thresholds, the IgG test was 91% sensitive and 99% specific for SARS-CoV-2, comparable to other highly sensitive spike-based assays (23, 41). The ROC analyses showed that further increases in sensitivity came at an unacceptable expense of specificity. Although the failure of the IgG test to detect spike-specific antibodies above threshold in ~10% of COVID-19 convalescents (at an average of 28 days post-symptom onset) may arise in part from technical limitations, it likely also reflects meaningful biological heterogeneity in the antibody response to natural infection (17, 42, 43). Our positive Conv cohort was composed solely of individuals characterized as having mild disease, with none requiring oxygen support. Recent work has shown that such individuals are more likely to seroconvert slowly and to have a lower overall antibody response (15, 19, 44–46).

The IgA test was considerably less sensitive (~70%) than the IgG at a threshold selected to provide 99% specificity (Fig 3a, c) in contrast to early reports of anti-S IgA assay that had higher sensitivity, but lower specificity than IgG (15, 19, 22, 25, 47). This is unlikely to be due to the delayed development of an IgA response relative to IgG, as the kinetics of IgA seroconversion has been shown to resemble, or even slightly precede that of IgG (15, 25). Rather, it may reflect the more rapid waning of serum IgA in convalescents (27). Concordantly, the IgA test appeared to detect a SARS-CoV-2 response in a higher percentage of samples from our small cohort of hospitalized patients (Fig 5). We also examined the possibility that, despite its lower sensitivity, the IgA test could be used to identify positives missed by the IgG test. We found that only 1% of the Conv cohort was positive for IgA alone, which was similar to the false-positive rate (Fig 2i, Table 4). We conclude that there is no added value to combining the IgG and IgA tests or using the latter for reflex testing to diagnose SARS-CoV-2 exposure. Spike-specific IgA may nevertheless be of use as a biomarker to help assess disease severity in acutely infected patients (15, 25).

The performance characteristics of the IgG test were used to assess its clinical utility at different levels of population seroprevalence (Table 5). Seroprevalence in New York City was ~20% at the end of April, (31, 48). Further, data from the New York City Department of Health and Mental Hygiene show that the seropositive rate in Bronx County to be 32.5% based on testing of over 17% of the county’s population (49). Seroprevalence at MMC, obtained during patient intake from April 4 through August 27, 2020, was 25.1% of 26,397 tests, using the Abbott SARS-CoV-2 IgG assay (obtained using the SlicerDicer function of MMC’s Epic Electronic Medical Record; E. Cadoff, personal communication). Under these conditions, the IgG test described herein has high positive and negative predictive value (PPV and NPV, respectively, of 97%). Accordingly, we translated it to a high-throughput, semi-automated platform in a CLIA-certified laboratory at MMC. A high degree of concordance was observed between the manual and automated tests and between the manual test and a microsphere immunoassay test developed by NYSDOH’s Wadsworth Laboratory. More comprehensive cross-validation efforts comparing the Einstein/MMC IgG test with several FDA-authorized commercial antibody tests and other tests currently in development are being conducted at MMC (Wolgast, L. et. al., manuscript in preparation). These findings strongly support the clinical deployment of our IgG test.

**Table 5:** Positive and negative predictive values of IgG test at different levels of seroprevalence.

| Prevalence<br>(n=100,000) | Sens.<sup>a</sup><br>(%) | Spec.<sup>b</sup><br>(%) | Positive tests /<br>COVID+<sup>c</sup> | Positive tests /<br>COVID-<sup>d</sup> | PPV<sup>e</sup><br>(%) | NPV<sup>f</sup><br>(%) |
| --- | --- | --- | --- | --- | --- | --- |
| 1% | 91 | 99 | 860 / 1,000 | 990 / 99,000 | 48% | 99.9% |
| 10% | 91 | 99 | 8,600 / 10,000 | 900 / 90,000 | 91% | 99% |
| 20% | 91 | 99 | 17,200 / 20,000 | 800 / 80,000 | 96% | 98% |
| 30% | 91 | 99 | 27,300 / 30,000 | 700 / 70,000 | 98% | 96% |

<sup>a</sup> Sensitivity at cutoff of 0.9
<sup>b</sup> Specificity at cutoff of 0.9
<sup>c</sup> Calculated number of positive tests in the group of true positives
<sup>d</sup> Calculated number of positive tests in the group of true negatives
<sup>e</sup> Positive predictive value: likelihood that a positive test predicts a true positive.
<sup>f</sup> Negative predictive value: likelihood that a negative test predicts a true negative.

Finally, we used independent experimental datasets from >200 convalescent sera to generate a logistic regression model for the accurate estimation of IgG titers from single absorbance values obtained with the IgG test. These findings expand the research and diagnostic utility of the Einstein/MMC IgG test without sacrificing its simplicity and throughput. Specifically, we believe that our test will help meet the need for quantitative serology engendered by the development and deployment of spike-based vaccines and convalescent plasma transfusion therapy for COVID-19 (23). Indeed, given the significant percentage of COVID-19 convalescents with low or negative serologic reactivity in this study, the rapid but accurate measurement of antibody levels in plasma will be crucial to vetting plasma collected from convalescent donors (50, 51). Further, the rapid measurement of serum IgG and/or IgA in a point-of-care setting may find utility in clinical decision-making, including patient selection for the administration of medications such as steroids or convalescent plasma (52, 53) to treat COVID-19.

## Data Availability

All data are available from the contact author, Kartik Chandran, upon request.

## Acknowledgments

We thank E. Gutierrez, E. Valencia, L. Polanco, and S. Diaz for laboratory management and technical assistance. We are grateful to J. McLellan for providing a plasmid expressing a stabilized SARS-CoV-2 spike ectodomain. We thank J. Achkar, M. Hawkins, R. Ostfeld, B. Rudolph, for their generous donation of control sera and J. LaFleur and other staff members of the Einstein Biorepository for assistance with the samples and associated metadata. We thank J. Wigren Byström for assistance with the Swedish hCoV patient samples. We thank members of the Diagnostic Immunology Laboratory at the Wadsworth Center/NYSDOH for their assistance with the cross-validation analysis. This work was supported by National Institutes of Health grants AI142777 (to K.C.); AI125462 (to J.R.L.), AI23654, AI143153, and 3UL1TR002556-04S1 (to L.P.); AI124753, AI13261, and AI134753 (to L.M.W.); AI145024 and the Einstein-Rockefeller-CUNY Center for AIDS Research (P30AI124414) (to S.C.A.). K.C. and J.R.L. were also supported by a COVID-19 Pilot Award from the Albert Einstein College of Medicine. L.P. was also supported by a grant from the G. Harold and Leila Y. Mathers Charitable Foundation. S.C.A. also acknowledges the Albert Einstein Macromolecular Therapeutics Development Facility. M.E.D. is a Latin American Fellow in the Biomedical Sciences, supported by the Pew Charitable Trusts. R.H.B.III. and R.J.M. were partially supported by NIH training grant 2T32GM007288-45 (Medical Scientist Training Program) at Albert Einstein College of Medicine. K.C. is a member of the scientific advisory board of Integrum Scientific LLC.

## Materials and Methods

### Patient Cohorts

Protocol approvals for patient sample acquisition were obtained by the Institutional Review Boards (IRB) of the Albert Einstein College of Medicine (Conv, Hosp, Ctrl cohorts, US samples in hCoV cohort) or Umeå University Hospital (Swedish samples in hCoV cohort).

#### Control (Ctrl) Cohorts

Patient sera samples collected prior to identification of the first case of COVID-19 in the United States.

#### Ctrl-2020

45 de-identified remnant sera from unique patients collected in January 2020.

#### Ctrl-Pre2020

171 de-identified remnant sera from unique patients, collected between October 1 and January 1 in 2008 to 2019 and stored in the Einstein Biorepository. These months were selected to enrich for samples from patients with non-COVID-19 respiratory viral illnesses. Samples were collected for a variety of studies, but those from studies that enrolled HIV-infected patients were excluded.

#### Human coronavirus (hCoV)

Remnant sera from patients with confirmed positive RT-qPCR tests for HCoV-229E, HCoV-OC43, HCoV-NL63, or HCoV-HKU1. Five sera were collected in January and early February 2020 and were identified from remnant sera in the MMC Pathology Laboratory. Another seventeen were from samples collected from patients in Umeå, Sweden in 2019-2020. All patient samples collected in 2020 were confirmed negative for SARS-CoV-2 by RT-qPCR.

#### Hospitalized (Hosp)

De-identified remnant sera from MMC inpatients who tested positive for COVID-19 by the SARS-CoV-2 swab molecular test. A total of 27 patients with sera samples collected at initial presentation in the Emergency Department or on days 0-1(early) and 7-10 days post admission (late) were selected for analysis. Clinical data indicating how long symptoms were present before admission to the hospital was also available for most patients and were used to redefine samples by days post-symptom onset (e.g. if a patient had a history of 5 day of symptoms, the specimen was treated as day 5 in this series).

#### Convalescent (Conv

De-identified samples from 197 healthy adult volunteers in Westchester County, NY who had previously recovered from COVID-19 were collected as indicated below. All patients had confirmed past SARS-CoV-2 infection with documented positive RT-qPCR. All patients were at least 14 days post resolution of symptoms (≥ 30 days post infection) at the time of collection. These sera were initially collected to screen potential convalescent plasma donors for a clinical trial on the efficacy of convalescent plasma for the treatment of COVID-19.

### Sample collection and handling

Conv and Hosp cohort sera were obtained by venipuncture (BD Vacutainer, Serum), centrifuged, aliquoted and stored at −80°C. Prior to analysis for anti-spike IgG and IgA antibodies, samples were heat-inactivated for 30 minutes at 56°C and stored at 4°C. Samples were handled under BSL-2 containment in accordance with a protocol approved by the Einstein institutional biosafety committee. Historical serum samples (Ctrl-2020 and Pre2020 and hCoV cohorts) were previously stored at −80°C. Aliquots were thawed, heat inactivated as above, and stored at 4°C prior to analysis.

### Protein Production and Purification

A pCAGGS plasmid encoding a mammalian codon-optimized, stabilized SARS-CoV-2 spike protein with C-terminal Twin Strep and 8X His Tags (gift from Jason McLellan, described in Wrapp et. al., 2020) was transiently transfected into ExpiCHO-S^TM^ cells (Gibco, Gaithersburg, MD #A29127) (0.8 µg DNA per mL of ExpiCHO-S^TM^ culture) as per the manufacturer’s instructions. Cells were incubated at 37°C for 1 day, then at 32°C in a shaking incubator (125 RPM with 8% CO2) and fed according to the manufacturer’s high titer protocol. Supernatant was harvested on day 12 by centrifugation at 3700xg for 20min, adjusted to pH 8 and dialyzed overnight at 4°C in Tris buffer (50 mM Tris HCl [pH 8.0], 250 mM NaCl). Supernatant was incubated with Ni-NTA resin for 2 hours at 4°C before resin was collected into a column and washed with Tris buffer plus 20 mM Imidazole. Spike protein was eluted with Tris buffer plus 250 mM Imidazole. Eluant was concentrated in an Amicon Ultra-15 100,000 NMWL Centrifugal Filter unit (Millipore Sigma, Burlington, MA #UFC9010) and buffer exchanged by dialysis into Tris buffer. Protein was aliquoted, flash frozen in liquid nitrogen and stored at −80°C. Protein quality was confirmed by analytical size exclusion chromatography using a Superose^TM^ 6 Increase 10/300 GL (Cytiva, Marlborough, MA) before and after flash freezing.

### Spike-specific IgG and IgA ELISA

Half-area ELISA plates (Corning #3690, Corning, NY) were incubated overnight at 4°C with 25 µL per well of 2 μg/mL of purified SARS-CoV-2 spike protein or S1 subunit from MERS (Sino Biological, Chesterbrook, PA 40069-V08H), or HKU-1 (Sino Biological 40602-V08H), 229E (Sino Biological 40601-V08H). Plates were washed three times with 120 µL per well 1× PBS-T (1× PBS, pH 7.4 + 0.1% (v/v) Tween-20) using a microplate washer (BioTek, Winooski, VT) before being blocked for 1h at 25°C with 150 μL per well of 1× PBS-T + 3% (v/v) milk (Bio-Rad #170-6404). Serum was serially diluted in 96-well non-tissue culture treated round-bottom plates (CELLTREAT, Pepperell, MA #22991) using 1× PBS-T + 1% (v/v) milk (1% milk PBS-T) as the diluent. Blocked ELISA plates were washed three times with 120 μL per well of 1× PBS-T, then 25 µL of diluted serum was added to wells in duplicate. Plates were incubated for 2h at 25°C before being washed three times with 120 μL per well of 1× PBS-T. Plates were incubated for 1h at 25°C with 25 µl of secondary antibody (1:3,000 in 1% milk PBS-T): goat anti-human IgG-Horseradish peroxidase (HRP) produced in goat (Invitrogen, Carlsbad, CA #31410) or anti-human IgA-HRP produced in goat (Millipore Sigma #A0295). Plates were washed as before, prior to development with 25 μL per well of ultra-TMB ELISA substrate solution at room temp (Thermo Scientific #34029). Plates were incubated in the dark for 5 min before quenching the reaction with 25 µL per well of 0.5 M sulfuric acid (Millipore Sigma #339741). Absorbance at 450 nm (A_450_) was measured using a Cytation 5 plate reader (BioTek).

### Clinical Pathology Lab ELISA

ELISA plates (Corning #3690) were coated with purified SARS-CoV-2 spike protein and washed and blocked as described above. Aliquots from the samples used for development of the research assay (see above) were diluted manually using 1% milk PBS-T as the diluent to obtain the targeted single dilution for IgG (1:1000) or IgA (1:200). Each of these was added in coated ELISA wells by the DSX automated ELISA system (Dynex Technologies; Chantilly VA) using disposable pipette tips. Incubations, washings, and addition of the anti-human IgG and IgA HRP conjugates, as well as the addition of substrate and stop solution was also accomplished using the automated system. Wash buffer and all reagents were identical to those used for the research assay described above. Reading was accomplished using a dual wavelength mode with the same test wavelength (450 nm) as the research assay and a reference wavelength of 620 nm.

### Nonlinear regression analysis

We used nonlinear least-squares analysis to fit a sigmoidal function (Eq: 1) to the experimental data (log_10_ IgG titer and A_450_ using a 1/1000 diluted serum; Fig 7):

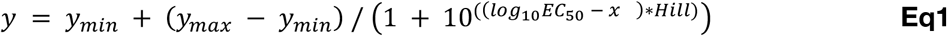

Where *y* corresponds to the absorbance; *y_min_* and y_*max*_ are the minimum and maximum absorbances respectively; *EC_50_* is the IgG titer that gives half of maximum absorbance *y_max_*; *Hill* describes the slope of the curve and; *x* is the log_10_ of the IgG titer.

To predict IgG titers for a given A_450_ value (measured using 1/1000 diluted serum) we first inferred A_450_ using the fitted sigmoidal model, for 10,000 IgG titers evenly spaced between the experimentally observed minimum and maximum IgG titers in our dataset. We then identified the closest inferred A_450_ to the queried A_450_ value and interpolated the corresponding IgG titer. We evaluated our nonlinear model by 10-fold cross-validation, where the original dataset is randomly partitioned into 10 equal size subsets, and one of the subsets serves as the testing set while the remaining nine subsets are used for training the non-linear model. This process is repeated 10 times, using subsets for testing and training each time to ensure that all data points in our dataset have been used once for testing. We iteratively (1,000 iterations) evaluated our nonlinear model by 10-fold cross-validation, computing the R^2^ value between the observed and predicted IgG titers at each iteration. Non-linear regression was performed using the scipy library (54).

